# Preliminary Analysis of the Melanoma Multimedia Educational programme for general practitioners on behalf of the Italian Melanoma Intergroup

**DOI:** 10.1101/2023.12.28.23300602

**Authors:** Federica Zamagni, Fabio Falcini, Serena Magi, Lauro Bucchi, Silvia Mancini, Rosa Vattiato, Emanuele Crocetti, Stefano Falcinelli, Claudio Feliciani, Maurizio Lombardo, Davide Melandri, Patrizia Re, Francesco Ricci, Rosanna Rita Satta, Sara Gandini, Ignazio Stanganelli, FAD MelaMEd Working Group

## Abstract

**Introduction:** According to the National Oncological Plan 2023-2027 on the importance of multidisciplinary and interactive e-learning training, the Italian Melanoma Intergroup (IMI) has developed MelaMEd (Melanoma Multimedia Education), a national project for general practitioners (GPs) on the prevention and detection of cutaneous melanoma through an online platform and an online course. MelaMEd enables participants to (1) recognize skin lesions that require specialist dermatological assessment, (2) select patients at high risk of melanoma and (3) be informed of the diagnosis and treatment pathway of patients with melanoma.

**Methods:** A free online platform and online course were developed and launched in June 2022. Before starting the course, enrolled participants fill out a pre-training questionnaire concerning the basic knowledge of the disease and the recognition and management of suspicious lesions. After the course, participants will fill out the same questionnaire again. The online course will end in December 2023. Here we present a preliminary analysis of the pre-training results (January 2023-July 2023). The data have been analyzed descriptively.

**Results:** So far, five healthcare centers have participated in the project for a total of 1320 participants. Of these, 298 compiled the pre-training questionnaire. Forty-seven percent of them were aged <40 years. Respondents were almost divided between GPs (47%) and resident GPs (48%). Among the theoretical questions, the “ABCDE” rule and “ugly duckling” sign are well known (96% and 91% of correct answers, respectively), but a lower percentage (68%) of respondents knows the “EFG” rule for the recognition of nodular melanomas and the statement of Breslow thickness (29%).

Regarding the series of clinical images of pigmented skin lesions and their management, the percentages rate of accuracy varied from 33% to 87%: melanoma (5 cases) ranges from 36% to 71%, melanocytic nevi (3 cases) from 33% to 84%, whereas the percentages rate of referral for dermatological evaluation varied from 44% to 99%. Melanoma cases referred to dermatologist ranges from 67% to 99%.

**Conclusions:** This preliminary analysis on pre-training questionnaire mainly showed a lack of knowledge of the two major points of melanoma diagnosis (EFG) and management (Breslow thickness), as well as a low rate of participants. We will compare the proportions of correct answers to the questionnaires before and after the course once available.

## Introduction

The rates of cutaneous melanoma have been rising rapidly over the past few decades in fair-skinned populations over the world. It is the sixth most frequently occurring cancer (after breast, colorectal, prostate, lung, and bladder cancers) and one of the 20 most frequent causes of cancer death melanoma in EU-27 countries (https://ecis.jrc.ec.europa.eu, accessed 15/05/2021). In the Italian population it is the second most frequent tumour in males under 50 years of age and the third most frequent in females under 50 years of age and the net survival is > 90% for both sexes also due to the introduction of target therapies and immune-checkpoint inhibitors.^1,2^

Although the great effort for primary prevention, the global burden of cutaneous melanoma has estimated to increase to 510,000 new cases (a roughly 50% increase) and to 96,000 deaths (a 68% increase) by 2040.^3^ Early detection or secondary prevention significantly improves morbidity and mortality.^4^

In the clinical practice a visual evaluation to detect a suspicious skin lesion is one of the most rapid and cost-effective methods as crucial point of secondary prevention. For these reason primary care physicians, i.e. general practitioners (GP) are commonly the first medical triage to select suspicious lesion or identify high-risk melanoma patients for dermatological evaluation. On the contrary, they may have a pivotal role in melanoma management in populations where dermatology access gaps exist.^5^ However, literature data showed most GP do not receive a comprehensive training in melanoma diagnosis and management and educational interventions are strongly recommended.^5-7^ A recent review conducted to collect data about previously reported skin cancer interventions for GP varied widely in design, including literature-based interventions, live teaching sessions, scheduled course time (range from 5 min to 24 months). Several training courses demonstrated improvements in skin cancer knowledge and competency but only a few revealed positive clinical practice changes by biopsy review or referral analysis.^8^ The most relevant successful results with practice change involved multimedia technologies with e learning, dermoscopy course and management according to guideline.^8^ With the rapid advancement of technology, multimedia digital education has become an essential part of modern medical community and e-learning tools are transforming knowledge and experience levels. According to the Italian Oncological Plan 2023-2027 planned by Health Ministry on the importance of multidisciplinary and interactive e-learning training, the Italian Melanoma Intergroup (IMI) has developed MelaMEd (Melanoma Multimedia Education) project, a national programme for GP on the prevention and detection of cutaneous melanoma through an online platform and an online course. MelaMEd enables participants to (1) recognize skin lesions that require specialist dermatological assessment, (2) select patients at high risk of melanoma and (3) be informed of the diagnosis and treatment pathway of patients with melanoma.

In Italy the effect formal training on GP was seldom evaluated,^9,10^ but a complete assessment of a multimedia educational programme has never been analyzed.

## Methods

The MelaMEd project is ongoing. A free online platform and online course (i.e. FAD) were developed and launched in June 2022. A detailed and completely description of the protocol has been published in this on-line edition of Dermatology Report.

Briefly, the protocol describes the following topics: 1) the “MelaMEd Programme”, 2) the primary and secondary objectives, 3) the study design (including the topics of asynchronous e-learning course entitled “Early diagnosis and management of the therapeutic diagnostic pathway of melanoma”, a comprehensive description of the structure and chapters of the “MelaMEd platform”, and the interactive function allowing the user to further explore any aspect of the e-learning course available in ‘switching’ modality with the virtual library of the MelaMEd platform), 4) a description of the phases of the study, 5) the inclusion and exclusion criteria, 6) the methodology (including pre-training and post-training questionnaires, questionnaires validation, and consent to the processing of personal data), and finally 7) statistical analysis.

For this preliminary analysis before starting the course, enrolled participants fill in a pre-training questionnaire concerning the basic knowledge of the disease and the recognition and management of suspicious lesions. After the course, participants will fill out the same questionnaire again. The deadline of the online course is planned in December 2023. Here we present a first preliminary analysis of results (January 2023-July 2023). The data have been analyzed descriptively, computing numbers and percentages of the answers to each question.

The centers included in this preliminary analysis are those whose participants had already completed the pre-training questionnaire on July 31^st^: local health authority of Romagna, local health authority of Parma, local health authority of Varese, local health authority of Sassari and IDI-IRCCS Dermatological Research Hospital of Rome. Others healthcare centers will have been involved until the end of 2023.

## Results

So far, five IMI centers have participated in the project for a total of 1320 enrolled participants. Of these, 298 compiled the pre-training questionnaire. **Table 1** shows the characteristics of total respondents. Forty-seven percent of them were aged <40 years. Respondents were almost divided between GPs (47%) and resident GPs (48%). **Table 2** and **Table 3** show the results of the pre-training questionnaire. Among the theoretical questions, the “ABCDE” and “ugly duckling” rules are well known (96% and 91% of correct answers, respectively), but a lower percentage (68%) of respondents knows the “EFG” rule for the recognition of nodular melanomas and the statement of Breslow thickness (29%). Among the 10 images, the percentages rate of accuracy varied from 33% to 87%. Specifically in the setting of melanoma (5 cases) ranges from 36% to 71%, melanocytic nevi (3 cases) from 33% to 84%. Seborrheic keratosis (1 case) and basal cell carcinoma (1 case) were correctly diagnosed by 78% and 87% respectively. Regarding the lesions recommended to dermatological evaluation the percentages rate of referral varied from 44% to 99%. Melanoma cases referred to dermatologist ranges from 67% to 99%.

**Table 1.**
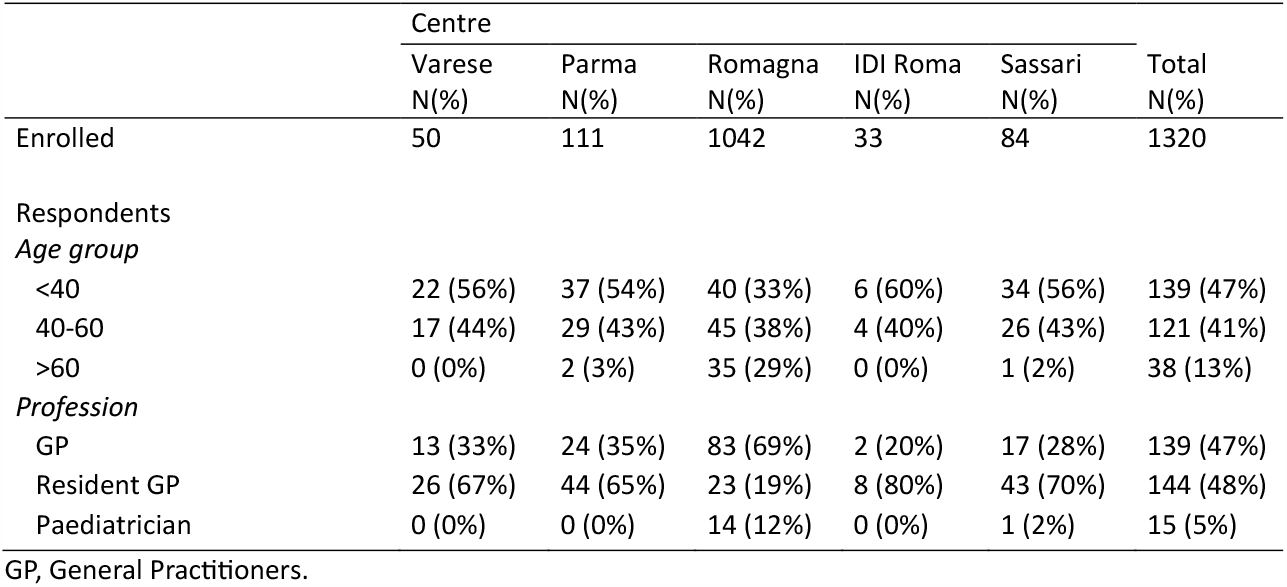
Characteristic of respondents, numbers and percentages by centre.

**Table 2.** Number and percentage of total correct answers to theoretical multiple-choice questions.

| Theroretical multiple choice questions | Correct answers |
| --- | --- |
|  | N (%) |
| Q1: Identify a risk factor for melanoma | 296 (99%) |
| Q2: choose what is needed for a complete visual examination of the skin | 275 (92%) |
| Q3: explain the acronym ABCDE | 287 (96%) |
| Q4: explain the EFG rule | 203 (68%) |
| Q5: explain what is meant by the "ugly duckling" sign | 271 (91%) |
| Q6: choose a true statement about dermoscopy | 187 (63%) |
| Q7: choose a true statement about Breslow thickness | 87 (29%) |

Q, question.

**Table 3.** Number and percentage of total correct answers to multiple-choice diagnosis for skin tumour images and number and percentages of referrals to dermatologist.

| Multiple-choice diagnosis for skin tumour images | Correct answers | Referrals to dermatologist |
| --- | --- | --- |
|  | N (%) | N (%) |
| I1: Thin melanoma | 121 (41%) | 278 (93%) |
| I2: Congenital melanocytic nevus | 97 (33%) | 219 (73%) |
| I3: Seborrheic keratosis | 232 (78%) | 188 (63%) |
| I4: Nodular melanoma | 212 (71%) | 287 (96%) |
| I5: Thick melanoma | 151 (51%) | 294 (99%) |
| I6: Melanocytic nevus | 249 (84%) | 185 (62%) |
| I7: Congenital melanocytic nevus | 232 (78%) | 130 (44%) |
| I8: Melanoma with regression | 173 (58%) | 293 (98%) |
| I9: Malignant lentigo | 108 (36%) | 201 (67%) |
| I10: Basal cell carcinoma | 258 (87%) | 295 (99%) |

I, image.

## Discussion

The need to improve the ability of GP in the evaluation of atypical melanocytic skin lesions can have a great impact in the secondary prevention of melanoma. Early detection of melanoma and the appropriate surgical treatment are all-important to ensure favorable outcomes in terms of morbidity and mortality. In addiction a comprehensive understanding of the primary and secondary prevention of cutaneous melanoma and a broad overview of diagnostic and therapeutic procedures may to be integrated in the community of primary-care health professionals, in particular GP.^8^

However inadequacy training for skin cancer management, variability in the identification of high risk patients, low diagnostic accuracy, unnecessary tests, inappropriate specialist referrals as well as economic, social and geographic barriers have been reported in the setting of primary-care health operators.^5-8^ Furthermore recent Italian data using a population cancer registries have suggested an overload primary care physician referral for a suspicious lesions for an increased patient presentation at dermatologic offices and have recommended a better GP triage in the referral patients.^11^

A recent review has been highlighted the importance and the effects of educational course on skin cancer detections. The most utilized form of the educational course was the short term, in-person, “live” delivery format, whereas multimedia and/or online format allows for the better dissemination of training to a wider audience. ^8^

The multimedia highly dynamic training of healthcare workers is strongly suggested, and the use of multimedia technologies to education with the widespread adoption of e-learning tools is being recommended.^12^

Furthermore, asynchronous e-learning solutions are made possible by web-based educational platforms, giving users the flexibility to finish their training at a time and method that suits them best.^8^

Brown et al showed improvements in skin cancer knowledge and expertise with the best results using multimedia technologies with e learning tools and dermoscopy course whereas poor results have been reported with short-term course without active interaction.^8^

In Italy the effects of educational course on melanoma are scarce and focused on diagnosis of melanoma or simulators (melanocytic nevi and non-melanoma skin cancer). Carli et al after training in a short-term course in a small group of participants (41 GP) showed significantly improvement of melanoma diagnostic accuracy and a reduction to benign lesions sent to dermatologists.^9^ Argenziano et al conducted a long term randomized study, comparing dermoscopy versus naked-eye evaluation arm, and showed that the use of dermoscopy improves the ability of GP to triage lesions suggestive of skin cancer without increasing the number of unnecessary expert consultations.^10^

The Melanoma Multimedia Education programme aims to provide physicians a global diagnostic and therapeutic multimedia tool on melanoma. This programme has an interactive function allowing the user to further explore any aspect of the e-learning course with the ‘switching’ modality of MelaMEd platform. This first preliminary analysis showed a limited presence of participants (298 out of 1320 participants) in the pre-training questionnaire and forty-seven percent of them were aged <40 years. This result confirm that e-learning interventions require access to, and familiarity with, web-based educational platforms as confirmed by the limitation of the use of web-based platform among older Italian.^13,14^ Regarding the theoretical questions, a low percentage (68%) of respondents knows the “EFG” rule and have a moderate knowledge about the fatal nodular melanoma. Nodular subtypes more commonly present as thick lesions; improved diagnostic accuracy of these is therefore critical and is significantly associated with poorer diagnostic accuracy. The knowledge of Breslow thickness (29%), a crucial prognostic histopathological feature for surgical and medical therapy, has been almost neglected. Among the images, the percentage rate of accuracy ranges from 33% to 87%. The skin lesions were mainly misclassified in the spectrum of melanocytic lesions, especially in the melanoma cases, ranging from 36% to 71%. Specifically, malignant lentigo was recognized correctly by 36% and but was referred to dermatologist by 67% of respondents, whereas thick melanoma and nodular melanoma were sent to dermatological evaluation by 99% and 96% respectively. Clinical diagnosis of malignant lentigo can be challenging due to overlapping features with benign lesions such as solar lentigo, pigmented actinic keratosis, and others. Nevertheless, despite the low diagnostic accuracy of melanoma cases, a consistent part of the respondents sent the malignant lesions to dermatologist. For this reason, it is important the selection of GP to send these lesions to dermatologist to perform clinical and dermoscopical analysis.

## Conclusions

The multimedia training with the use of digital tools to medical education has been increasing with the fast and growing international spreading in the teaching and e learning processes.^12^

MelaMEd programme allows for providing physicians a wide diagnostic and therapeutic multimedia e learning tool on melanoma. This first preliminary analysis showed mainly a lack of knowledge of the two major points of melanoma diagnosis (EFG) and management (Breslow thickness). In the management of pigmented skin lesions, an improvement on diagnostic accuracy can be recommended, confirming the importance of multimedia training on melanoma management among primary healthcare area. In addition, the low rate of participants suggests the need for a better process of acquiring digital knowledge and skills among medical community. Finally, we will compare the proportions of correct answers to the questionnaires before and after the course once available, evaluating the impact of this complete multimedia educational programme.

## Data Availability

All data produced in the present study are available upon reasonable request to the authors

## Acknowledgments

The European Institute of Oncology, Milan, Italy is partially supported by the Italian Ministry of Health with Ricerca Corrente and 5×1,000 funds

## Working Group

The FAD MelaMEd Working Group includes:

Maria Antonietta Pizzichetta (Aviano, Trieste), Stefania Stucci, Marco Tucci (Bari), Vincenzo De Giorgi, Daniela Massi (Firenze), Paola Ghiorzo, Cesare Massone, Enrica Tanda (Genova), Francesco de Rosa, Matelda Medri (Meldola), Roberto Patuzzo (Milano), Corrado Caracò, Marco Palla (Napoli), Alessio Fabozzi (Padova), Mario Mandalà (Perugia), Riccardo Marconcini (Pisa), Paolo Fava, Elena Marra, Pietro Quaglino, Simone Ribero (Torino) for the development of educational contents.

Anna Fanti, Fabiana Perrone, Rocco Tortorella (Parma), Barbara Andrea, Paolo Parente, Andrea Silenzi (Roma), Emanuela Manzato, Carlo Somenzi (Romagna), Gabriele Biondi, Chiara Musio, Giuseppe Palmieri (Sassari), Marco Paolo Donadini, Giovanna Scienza (Varese), for the organization at the local health unit level.

## Supplementary materials

Raw data are accessible at the link: **the link will be available as soon as possible**

## References

1. Bucchi L, Mancini S, Crocetti E, et al. Mid-term trends and recent birth-cohort-dependent changes in incidence rates of cutaneous malignant melanoma in Italy. Int J Cancer. 2021 Feb 15;148(4):835–844.

2. Zamagni F, Bucchi L, Mancini S, et al AIRTUM Working Group. The relative contribution of the decreasing trend in tumour thickness to the 2010s increase in net survival from cutaneous malignant melanoma in Italy: a population-based investigation. Br J Dermatol. 2022 Jul;187(1):52–63.

3. Arnold M, Singh D, Laversanne M, et al. Global Burden of Cutaneous Melanoma in 2020 and Projections to 2040. JAMA Dermatol. 2022 May 1;158(5):495–503.

4. AM Forsea. Melanoma Epidemiology and Early Detection in Europe: Diversity and Disparities. Dermatol Pract Concept. 2020 Jul;10(3):e2020033.

5. Ezeonwu MC. Specialty-care access for community health clinic patients: processes and barriers. J Multidiscip Healthc. 2018;11:109–19.

6. Fleischer AB Jr., Herbert CR, Feldman SR, O’Brien F. Diagnosis of skin disease by non-dermatologists. Am J Manag Care. 2000;6(10):1149–56.

7. Kirsner RS, Muhkerjee S, Federman DG. Skin cancer screening in primary care: prevalence and barriers. J Am Acad Dermatol. 1999;41(4):564–6.

8. Brown AE, Najmi M, Duke T, et al. Skin Cancer Education Interventions for Primary Care Providers: A Scoping Review. J Gen Intern Med. 2022 Jul;37(9):2267–2279.

9. Carli P, De Giorgi V, Crocetti E, et al. Diagnostic and referral accuracy of family doctors in melanoma screening: effect of a short formal training. Eur J Cancer Prev 2005;14:51–5.

10. Argenziano G, Puig S, Zalaudek I et al. Dermoscopy improves accuracy of primary care physicians to triage lesions suggestive of skin cancer. J Clin Oncol. 2006 Apr 20;24(12):1877–82.

11. L Bucchi, S Mancini, F Zamagni et al. Patient presentation, skin biopsy utilization and cutaneous malignant melanoma incidence and mortality in northern Italy: Trends and correlations. J Eur Acad Dermatol Venereol. 2022 Sep 30.

12. Abdulrahaman MD, Faruk N, Oloyede AA, et al. Multimedia tools in the teaching and learning processes: A systematic review. Heliyon. 2020 Nov 2;6(11):e05312.

13. Civitelli G, Tarsitani G, Rinaldi A, Marceca M. Medical education: an Italian contribution to the discussion on global health education. Global Health. 2020 Apr 8;16(1):30.

14. Marceglia S, Balestra G, Bottrighi A, et al. Developing the Digital Healthcare Workforce in Italy: The SIBIM Experience. Stud Health Technol Inform. 2022 Aug 31;298:46–50.

